# Establishing Clinically Significant Patient-reported Outcomes for Diverticular Disease – A Prospective Cohort Study

**DOI:** 10.1101/2020.02.21.20026427

**Authors:** Sara Khor, David R. Flum, Lisa L. Strate, Mariam N. Hantouli, Heather H. Harris, Danielle C. Lavallee, Brennan MR Spiegel, Giana H. Davidson

## Abstract

**Background & Aims:** Diverticular disease can undermine health-related quality of life (HRQoL). The diverticulitis quality of life (DV-QOL) instrument was designed and validated to measure the patient-reported burden of diverticular disease. However, in order for the DV-QOL to capture longitudinal outcomes, values reflecting meaningful improvement (i.e., minimal clinically important difference (MCID)) and the patient acceptable symptom state (PASS) need to be established. We sought to establish the MCID and PASS of the DV-QOL, and describe the characteristics of those with DV-QOL above the PASS threshold.

**Methods:** We performed a prospective cohort study of adults with diverticular disease from 7 centers in Washington and California (2016-2018). Patients were surveyed at baseline, then quarterly up to 30 months. To determine the PASS for DV-QOL, we used an anchor-based approach and performed receiver operating characteristic analyses using baseline data. A range of MCID values were calculated using distribution-based and anchor-based approaches.

**Results:** The study included 177 patients (mean age 57, 43% female). A PASS threshold of 3.2/10 distinguished between those with and without HRQoL-impacting diverticulitis with acceptable accuracy (area under the curve (AUC) 0.76). A change of 2.2 points in the DV-QOL was the most appropriate MCID: above the measurement error and corresponding to patient perception of importance of change (AUC 0.70). Patients with DV-QOL≥PASS were more often male, younger, had Medicaid, less educated, and had more serious episodes of diverticulitis.

**Conclusions:** Our study is the first to define MCID and PASS for DV-QOL. These thresholds are critical for measuring the impact of diverticular disease and the evaluation of treatment effectiveness.

## Background

Two to three million Americans experience an episode of acute diverticulitis each year, resulting in over 370,000 emergency department (ED) visits, 200,000 hospital admissions, and almost $5.5 billion in total health care expenditures.^1^ One in four patients will have recurrent episodes of diverticulitis, and historically, patients with more than a few episodes (ranging from 1-3, based on risk) were recommended to have an elective colectomy.^2,3^ In 2015, the American Society of Colorectal Surgeons (ASCRS) proposed that an “episode count” indication for elective colon resection be replaced by an assessment of the impact of diverticulitis on patient-reported health-related quality of life (HRQoL).^4^ The evolution in the ASCRS guideline demonstrates a recognition that some patients who have recovered from an acute episode of diverticulitis often have ongoing gastrointestinal symptoms and psychosocial distress after recovery from an episode.^5^ This emphasizes the importance of a reliable metric to understand the impact of diverticulitis on patient-reported symptoms and HRQoL domains over time.

In 2015, Spiegel et al. developed the first diverticulitis-specific HRQoL measure (DV-QOL),^6^ a 17-question survey of patient-reported burden of disease focused on those with uncomplicated diverticular disease. The DV-QOL includes questions about both intestinal and extra-intestinal symptoms, behavior changes related to the disease, and disease-specific cognitions and emotions. Each question of the DV-QOL is worded so that questions attribute symptoms specific to diverticular disease, unlike more generic patient-reported outcome (PRO) measures, such as the Gastrointestinal Quality of Life Index (GIQLI) or the 36-Item Short Form Health Survey questionnaire (SF-36).^7^ While generic measures are useful for comparing outcomes across different populations and programs, disease-specific tools have been suggested as a way to capture specific concerns for patients with particular conditions and may be helpful for measuring clinically important changes with treatment.^8^ This is particularly important for investigators and clinicians who need tools that include specific features of a particular disease and are responsive to treatments and variations in health status over time.

However, important questions remain regarding the use of the DV-QOL for clinical application, e.g., establishing the change in score that is clinically relevant and determining the lowest score below which patients consider themselves well. The minimum clinically important difference (MCID) and the patient acceptable symptom state (PASS) are values that reflect meaningful improvement and patient satisfaction, respectively.^9,10^ Determining the MCID and the PASS of the DV-QOL is critical for interpreting the scores of both individual patients and populations undergoing treatment and is essential to support treatment guidelines. The goals of this study were to establish the PASS and MCID of the DV-QOL and to explore additional characteristics associated with patients reporting more severe disease on the DV-QOL.

## Methods

### Study Design and Population

This was a prospective cohort study with patients recruited between April 1, 2016, and November 30, 2018, from the Diverticulitis Evaluation of Patient Burden, Utilization, and Trajectory (DEBUT) study.^11^ Patients with a history of acute diverticulitis were recruited from different clinic environments (EDs, surgery clinics, and gastrointestinal clinics) at seven medical centers in California and Washington and at different stages of disease (e.g., recent diagnoses or those with recurring episodes). Adult patients were recruited if they had a computed tomography (CT) scan-confirmed report or a physician-confirmed diagnosis of diverticular disease. Patients who had a prior colon resection, used a medical proxy for decisions about care, or were non-English speaking were excluded. Patients were predominantly recruited remotely via emails and letters. Of the 591 patients approached for the study, 214 (36%) agreed to participate; of these, 177 (83%) returned a completed baseline survey. Participants were surveyed at baseline and every 3 months via a multi-modal approach that included outreach by phone, mail, and email with options to complete the survey on paper or online. At the time of this analysis, the longest follow-up was 30 months.

The baseline survey collected information related to patient demographics (i.e., age, sex, race, ethnicity, marital status, education, insurance, job requiring physical activity, income), smoking status, diverticulitis history (i.e., years of disease, episodes of disease, timing and severity of the last episode), and recruitment site type.

### Outcome Measure

The DV-QOL is a 17-item questionnaire designed to assess patients’ disease-targeted HRQoL.^6^ It combines four domains relevant to the HRQoL of patients with diverticulitis: 1) physical symptoms, 2) concerns, 3) emotions, and 4) behavioral changes (see eTable 1 in the supplementary materials for items in each domain) and takes approximately 10-15 minutes to complete. The total DV-QOL score is reported on a 0-10 scale, with 0 indicating the lowest symptom burden (best HRQoL) and 10 the highest burden (worst HRQoL). The DV-QOL was collected at baseline and quarterly thereafter.

**Table 1:** Approaches used for determining the PASS and MCID thresholds of DV-QOL

| Approach |  | Description |
| --- | --- | --- |
| **PASS<sup>a</sup>** |  |  |
| **Anchor-based** | **ROC<sup>b</sup> curve-derived** | ROC curves were computed for each DV-QOL cutoff value. The corresponding ROC curve for each cutoff value represented the relation between the proportion of patients who were correctly classified in the acceptable group (sensitivity) versus the proportion of patients who were incorrectly classified in the acceptable group (specificity). The acceptable threshold is the value that provides the best balance between sensitivity and specificity, representing the lowest overall misclassification. |
| **MCID<sup>c</sup>** |  |  |
| **Anchor-based** | **1. Average change** | The mean DV-QOL change score<sup>d</sup> of the patients who improved |
|  | **2. Change difference** | The difference in the mean change scores between the patients who improved and did not improved |
|  | **3. ROC curve-derived** | The change score with even sensitivity and specificity based on the ROC curves, which was used to distinguish between patients who improved and those who did not. (See above for more detail.) |
| **Distribution-based** | **4. Half a standard deviation (SD)** | Half the SD of the DV-QOL scores at Q<sub>max</sub><sup>e</sup> |

<sup>a</sup>PASS, patient acceptable symptom state
<sup>b</sup>ROC, receiver operating characteristic
<sup>c</sup>MCID, minimum clinically important difference
<sup>d</sup>The score in the quarter with the highest DV-QOL score (Q<sub>max</sub>) minus the score in the subsequent quarter. A positive change score indicates improvement, and negative change represents deterioration in QoL.
<sup>e</sup>Q<sub>max</sub>, Quarter with the highest DV-QOL score

### Anchors

An anchor question asked about an individual’s HRQoL improvement over time: “Do you feel that your digestive health has improved over the last 3 months?” Patients could answer “Yes,” “No,” or “Not sure.” These answers were dichotomized into 1 = Improved (“Yes”) and 0 = Not improved (“No” or “Not sure”). Patients were asked to complete this anchor question at all time-points following the baseline survey.

An additional anchor question collected information regarding satisfaction with current digestive quality of life. At baseline and at each follow-up time-point, patients were asked, “Are you content with your digestive quality of life today?” Patients could answer “Yes,” “No,” or “Not sure.” These answers were further dichotomized into 1 = acceptable (“Yes”) and 0 = unacceptable (“No” or “Not sure”). Only a subset of patients (n=120/177, 68%) received this question at baseline.

### Patient Acceptable Symptom State (PASS)

An anchor to self-identified satisfaction was used to determine the DV-QOL threshold for the PASS that best differentiated patients who were currently content with their digestive HRQoL (acceptable) versus those who were not (unacceptable). We performed the receiver operating characteristic (ROC) analyses using baseline data (Table 1). The acceptable threshold for PASS is the value that provides the best balance between sensitivity and specificity, representing the lowest overall misclassification. The area under the curve (AUC) was also computed for each DV-QOL threshold, representing the probability of correctly differentiating between an acceptable and an unacceptable state. An AUC of 0.7 to 0.8 is considered acceptable, and an AUC of 0.8 or above excellent.^12^

### Minimum Clinically Important Difference (MCID)

To calculate the MCID, we used data from the quarter with the highest DV-QOL score (Q_max_) that was not the final quarter of the study participation. Only patients who had completed two consecutive quarters of the DV-QOL questionnaire and had a DV-QOL score at Q_max_ ≥ the PASS were included. Given the heterogeneity of disease severity of our cohort, this inclusion criterion was selected to capture meaningful improvement among those with HRQoL-limiting diverticular disease. Three anchor-based approaches (average change, change difference, and ROC-derived) and one distribution-based approach (0.5 standard deviation) were used to determine the MCIDs;^13,14^ these are outlined in Table 1. The anchor-based MCIDs were derived using the anchor question about HRQoL improvement. The final MCID value was chosen based on the fulfillment of two criteria: it must be at least greater than the measurement error (distribution-based), and it must correspond to the patient perception of importance of change (anchor-based).

### Statistical Analysis

We described the patient characteristics and disease history among patients with baseline DV-QOL below and above the PASS value to examine how the two groups differed in these factors. Univariate analysis was performed with respect to each variable using χ2 or the Student t test for categorical and continuous variables, respectively. We also examined the associations of HRQoL-limiting disease (i.e., DV-QOL above PASS) with other PROs at baseline, including the Patient-Reported Outcomes Measurement Information System (PROMIS) scores, self-reported chronic sickness, and work and activity impairment measures. The PROMIS global health instrument assesses an individual’s generic mental health (four items) and physical health (four items) domains.^15,16^ The total raw scores are translated into standardized T-scores, ranging from 0 (worst) to 100 (best), with a mean of 50 and a standard deviation (SD) of 10. Self-reported chronic sickness was measured by the question, “Do you feel sick all of the time?”^17^ The Work Productivity and Activity Impairment questionnaire is an instrument to measure impairments in both paid and unpaid work due to health problems.^18^ We expected patients with HRQoL-limiting disease to have lower (worse) PROMIS physical and mental health scores, to be more likely to feel chronically sick, and to report higher work and activity impairment. Variables were considered significant if the final p-value was <.05. Data analysis was performed using Stata version 14 (Stata-Corp). This study received IRB approval for all of the study locations.

## Results

Among 177 patients who filled out a baseline survey (43% female, mean age 57 years (SD 13)), 81 (46%) were recruited in EDs, 76 (43%) from surgical clinics, and 20 (11%) from gastrointestinal clinics. Table 2 shows patient characteristics at baseline. In this patient population, 79% were white, 51% had private insurance, and 50% were currently employed. Time since diverticulitis diagnosis varied greatly, with a median of 1.7 years (interquartile range (IQR) 0.2-7.5). The median baseline DV-QOL total score was 3.9 (IQR 2.2-5.6) out of 10 (Table 3). The domain related to concerns had the worst (highest) score (median 5.0; IQR 2.5-7.5). There were 31 (18%) patients who reported feeling chronically sick; median PROMIS physical and mental health summary scores were 45 and 48, respectively, both within one standard deviation of the standardized mean score of 50. Median percentage of work and activity impairment due to heath was 20% (IQR 1-30%) and 30% (IQR 0-70%), respectively.

**Table 2:**
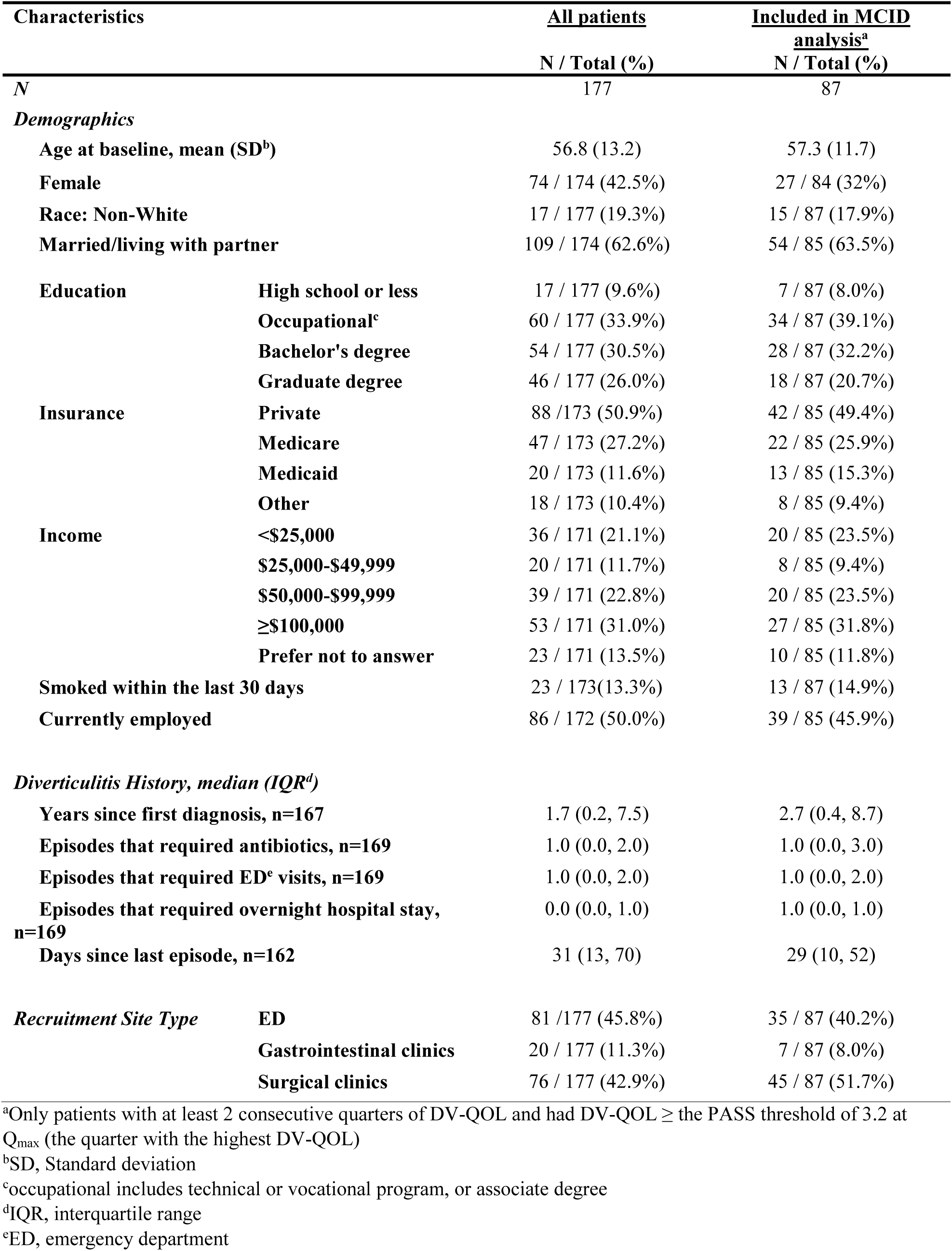
Patient baseline characteristics

**Table 3:**
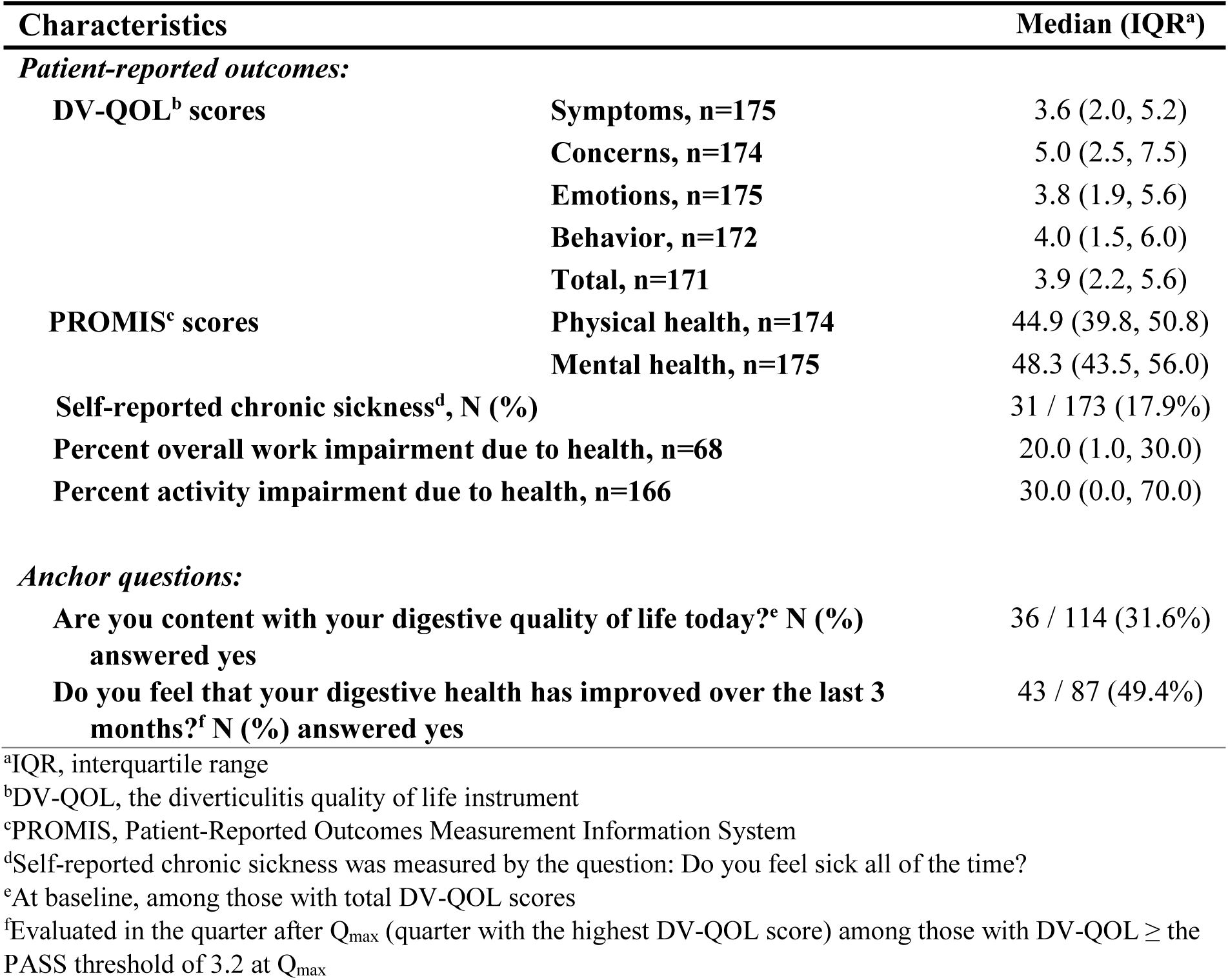
Patient-reported outcomes and anchor questions at baseline

### Establishing the Patient Acceptable Symptom State (PASS)

The anchor question regarding self-reported satisfaction with digestive HRQoL at baseline was answered by 114 patients. Of those, 36 (32%) reported they were content with their digestive HRQoL. DV-QOL was significantly higher among those who reported discontent (mean 4.89; SD 1.82) compared with those who were content (mean 2.55; SD 1.98). The DV-QOL cutoff value of 3.2 out of 10 represented the threshold that provided the best balance between sensitivity and specificity (AUC of the ROC curve 0.76; sensitivity 80.8%; specificity 72.2%) (eFigure 1).

### Establishing the MCID

In the study population, 132 patients had at least two consecutive quarters of data. Of those, 87 (66%) had a DV-QOL score at Q_max_ ≥ 3.2, the PASS threshold. Baseline characteristics of these patients were similar to those in the baseline analysis (Table 2). Mean DV-QOL scores were 5.56 (SD 1.4) and 2.80 (SD 1.9) for Q_max_ and the quarter after Q_max_, respectively. There were 43 (49.4%) patients who answered “Yes” to the improvement anchor question the quarter after Q_max_, and their mean DV-QOL change score was significantly higher than those who answered “No” or “Not Sure” to the improvement question (mean change score 3.80 (SD 2.1) versus 1.73 (SD 1.8); p<0.001). Each MCID calculation method yielded a different threshold. The average change method yielded the largest threshold (3.80). The change difference method and ROC curve methodology yielded thresholds of 2.07 and 2.20, respectively. The distribution-based method yielded the smallest threshold (0.72). Applying our choice criteria, the ROC curve-derived value of 2.20 appeared to be the most appropriate MCID because it was above the measurement error and close to the change difference. At the cutoff of 2.20, the AUC was 0.7 (sensitivity 74.4%; specificity 65.9%; eFigure 2).

### Variables associated with clinical significance

Table 4 shows the patient characteristics and diverticulitis history by HRQoL-limiting disease (defined by DV-QOL scores above the PASS threshold). Univariate analysis showed that patients who had HRQoL-limiting diverticulitis were younger, more often male, received less education, more often had Medicaid as their primary insurance, and had more episodes of diverticulitis attacks that required an ED visit or overnight hospital stays. HRQoL-limiting diverticulitis was significantly associated with worse PROMIS global health scores in both the physical health and mental health domains, a feeling of being “always sick,” and higher work and activity impairment (Table 5).

**Table 4:** Patient baseline characteristics by DV-QOL below/above PASS threshold (DV-QOL=3.2).

| Characteristics |  | DV-QOL<sup>a</sup> <3.2<br>N(%) | DV-QOL ≥3.2<br>N(%) | p-value |
| --- | --- | --- | --- | --- |
| N |  | 69 | 102 |  |
| Age at baseline, mean (SD<sup>b</sup>) |  | 59.2 (12.2) | 54.9 (13.3) | 0.040 |
| Female |  | 38/69 (55%) | 34/99 (34%) | 0.008 |
| Race Non-White |  | 10/69 (14%) | 20/102 (20%) | 0.33 |
| Married/living with partner |  | 42/69 (61%) | 63/99 (64%) | 0.72 |
| Education | High school or less | 5/69 (7.2%) | 11/102 (10.8%) | 0.018 |
|  | Occupational<sup>c</sup> | 15/69 (21.7%) | 41/102 (40.2%) |  |
|  | Bachelor's degree | 23/69 (33.3%) | 30/102 (29.4%) |  |
|  | Graduate degree | 26/69 (37.7%) | 20/102 (19.6%) |  |
| Insurance | Private | 34/68 (50%) | 51/99 (52%) | 0.010 |
|  | Medicare | 25/68 (37%) | 20/99 (20%) |  |
|  | Medicaid | 2/68 (3%) | 17/99 (17%) |  |
|  | Other | 7/68 (10%) | 11/99 (11%) |  |
| Income | <\$25000 | 10/65 (15.4%) | 23/101 (22.8%) | 0.38 |
| | \$25,000-\$49,999 | 6/65 (9.2%) | 14/101 (13.9%) | |
| | \$50,000-\$99,999 | 18/65 (27.7%) | 20/101 (19.8%) | |
| | ≥\$100,000 | 24/65 (36.9%) | 29/101 (28.7%) | |
|  | Prefer not to answer | 7/65 (10.8%) | 15/101 (14.9%) |  |
| Smoked within the last 30 days |  | 7/66 (10.6%) | 15/101 (14.9%) | 0.43 |
| Currently employed |  | 34/66 (51.5%) | 51/100 (51.0%) | 0.95 |
| *Diverticulitis history, median (IQR<sup>d</sup>):* |  |  |  |  |
| Years since first diagnosis |  | 1.7 (0.3, 8.3) | 1.8 (0.1, 7.0) | 0.50 |
| Episodes that required antibiotics |  | 1.0 (0.0, 3.0) | 1.0 (0.0, 2.0) | 0.71 |
| Episodes that required ED<sup>e</sup> visits |  | 0.0 (0.0, 1.0) | 1.0 (0.0, 2.0) | <0.001 |
| Episodes that required overnight hospital stay |  | 0.0 (0.0, 1.0) | 1.0 (0.0, 1.0) | 0.034 |
| Days since last episode |  | 54 (24, 148) | 19 (5, 49) | <0.001 |

<sup>a</sup>DV-QOL, the diverticulitis quality of life instrument
<sup>b</sup>SD, Standard deviation
<sup>c</sup>occupational includes technical or vocational program, or associate degree
<sup>d</sup>IQR, interquartile range
<sup>e</sup>ED, emergency department

**Table 5:** Patient-reported outcomes by DV-QOL below/above PASS threshold (DV-QOL=3.2).

| **Characteristics** |  | **DV-QOL<sup>a</sup> <3.2**<br>**Median (IQR<sup>b</sup>)** | **DV-QOL≥3.2**<br>**Median (IQR)** | **p-value** |
| --- | --- | --- | --- | --- |
| **N** |  | 69 | 102 |  |
| **PROMIS<sup>c</sup> scores** | **Physical health** | 50.8 (44.9, 54.1) | 39.8 (37.4, 47.7) | <0.001 |
|  | **Mental health** | 53.3 (48.3, 56.0) | 45.8 (41.1, 50.8) | <0.001 |
| **Self-reported chronic sickness<sup>d</sup>, n (%)** |  | 2/66 (3.0%) | 29/101 (28.7%) | <0.001 |
| **Percent overall work impairment due to health** |  | 0.2 (0.0, 27.8) | 23.6 (10.0, 39.7) | 0.002 |
| **Percent activity impairment due to health** |  | 0.0 (0.0, 10.0) | 70.0 (30.0, 80.0) | <0.001 |

<sup>a</sup>DV-QOL, the diverticulitis quality of life instrument
<sup>b</sup>IQR, interquartile range
<sup>c</sup>PROMIS, Patient-Reported Outcomes Measurement Information System
<sup>d</sup>Self-reported chronic sickness was measured by the question: Do you feel sick all of the time?

## Discussion

As clinical guidelines for elective colectomy in patients with diverticular disease focus on more personalized treatment related to disease burden and HRQoL limitation,^4^ instruments like the DV-QOL can play a critical role in comparative effectiveness research. Ultimately, clinical shared decision-making discussions for patients with diverticular disease may be enhanced by incorporating data that includes disease-specific PRO measures. Defining meaningful change and HRQoL-limiting state in a PRO score can help inform how well a treatment is working versus when new treatment approaches are needed. Our study is the first to define the MCID and PASS for the DV-QOL. Using prospectively collected data from a heterogeneous cohort of patients with diverticular disease, we found a score of 3.2 out of 10 distinguished between those with and without HRQoL-impacting diverticulitis, suggesting this PASS score as a threshold. We found a change of 2.2 points in the DV-QOL to be the most appropriate MCID because it was above the measurement error and corresponded to the patient perception of importance of change. These thresholds are critical for assessing burden of disease and measuring the impact of interventions in subpopulations of interest. We also identified patient characteristics at baseline associated with HRQoL-limiting disease (DV-QOL≥PASS), including male, younger age, less education, Medicaid status, and more episodes of diverticulitis that required an ED visit or overnight hospital stay. These factors may be considered in treatment counseling and when comparing the proportion of patients achieving the PASS between two or more treatment groups.

There is growing interest in using PRO measures in clinical practice, research, and policy. Instruments specific to different diseases or populations, rather than generic measures, are critical for identifying important concerns of patients with particular conditions and for measuring small, clinically important changes from specific treatments.^8^ Gastrointestinal system-specific instruments are becoming more widely used, such as the GIQLI and the PROMIS gastrointestinal (GI) symptom scales,^7,19^ but they are not disease-specific instruments for diverticular diseases. The disease-specific DV-QOL was developed through focus groups, literature search, and cognitive briefings with patients who had diverticular disease, capturing important experiences of illness using patients’ own words. The GIQLI and PROMIS GI have important overlap with the DV-QOL; for example, all three have questions about symptoms (e.g., abdominal pain, fullness and bloating in the abdomen, nausea, diarrhea, constipation), and the GIQLI and DV-QOL both have questions about the ability to complete normal daily activities (e.g., work), food restrictions, anxiety, and frustration. However, there are also some important differences. The DV-QOL is specific to diverticulitis, with each question attributing symptoms to the disease of interest as opposed to overall well-being. It also evaluates diverticulitis-specific patient concerns, such as whether diverticulitis might flare up or get worse at any time, or whether people might be looking down at them because of their diverticulitis symptoms. Questions in this domain are not assessed in the GIQLI nor the PROMIS GI. Our study found that the questions regarding condition-specific concerns received the highest (worst) scores compared with questions in other HRQoL domains, suggesting that the disease-specific concerns are an important domain of HRQoL for these patients. As a result of these differences, the DV-QOL is likely to be more sensitive to within-subject changes than generic measures,^8,20^ a hypothesis that is being tested in the currently running COSMID trial (PCORI #PCS-2018C1-10949).

We found characteristics associated with DV-QOL above PASS (DV-QOL≥3.2) at baseline that may be important to consider in future studies aiming to use the achievement of PASS as an outcome. These factors included male sex, younger age, less education, Medicaid status, and more episodes of diverticulitis attacks that required an ED visit or overnight hospital stay. Since our goal was to describe the differences in the population at baseline between those with high vs. low DV-QOL and not to predict future outcomes, we did not attempt to do a multivariable analysis. Future studies could explore the association between baseline characteristics and future outcomes. The PASS threshold in our study was also able to reasonably differentiate between patients with lower versus higher generic HRQoL (PROMIS scores), patients who were feeling sick all the time versus not, and those who experienced high versus low work and activity impairment because of their health. While we do not expect perfect correlation between DV-QOL and these external PRO measures, the general alignment (both in terms of direction and magnitude) of the DV-QOL-limiting state with other PRO measures suggests that the anchor-based DV-QOL threshold of 3.2 is a reasonable PASS value at baseline.

This study has important limitations. MCID and PASS thresholds were generated using data from English-speaking patients from California and Washington state, where 79% of the patients were white and 51% had private insurance, which limits the generalizability of our results.

Moreover, as a pragmatic observation study, our cohort reflects the heterogeneity of the population with diverticular disease and our study cohort consisted of patients with both severe and mild disease recruited from EDs and clinicians’ offices. The physician-confirmed diagnosis of diverticular disease relied on usual care, which may have included a CT scan, barium study, and/or colonoscopy. There may be differences in the MCID and PASS within subgroups that could not be evaluated in this cohort and this is critical to explore in future work. This study also combined patients receiving treatment at different stages of disease (e.g., 42 (24%) were diagnosed within 2 months of baseline, and 23 (13%) had elective surgery), although it is unclear how treatment or disease history will affect the MCID or the PASS thresholds. It is also possible that some patients had other GI comorbid illnesses that may have affected their DV-QOL scores that were not captured in this study. Furthermore, we did not randomize the order of the PRO measures in our surveys. The order of measurements may affect responses by potentially priming questions related to similar domains based on answers to the first questions about that domain.

Because DV-QOL questions were always asked first, we do not expect any impact on the MCID and PASS thresholds, but the order may have affected the answers to the PROMIS and work and activity impairment questions. Moreover, the determination of an MCID and PASS threshold using the ROC curve approach is subjective and determined by assessing the impact of the value on sensitivity and specificity, based on anchor questions. While balancing sensitivity and specificity is a widely used and conventional approach for MCID and PASS determination, some clinicians may favor thresholds that can identify patients with meaningful change or HRQoL-limiting disease with greater confidence (higher specificity) versus ruling out non-meaningful change or non-HRQoL-limiting disease (higher sensitivity). Nevertheless, the ROC-derived thresholds in our study had good sensitivity and specificity (range 68-82), and the MCID derived from the ROC method corresponds to the change difference between those who reported improvement and those who did not. Lastly, there is no gold standard “anchor question” to calculate the MCID and PASS thresholds. It may be the case that different MCID and PASS thresholds would have been calculated had different anchor questions been applied or if different choices of answers were provided. It is important to note that the other commonly used approach for determining MCID (0.5 SD) fell within the parameters determined by the anchor question methodology.

## Conclusions

People with diverticular disease experience a range of symptoms that may affect social and emotional health. Measuring the burden of diverticular disease is increasingly important for clinical decision making and evaluations of treatment effectiveness. This study expands upon existing work by establishing the MCID and PASS for the disease-specific DV-QOL, measures that are critical for the interpretation of PROs in individuals with diverticular disease.

## Data Availability

The DEBUT participating cohorts’ data for this analysis are not publicly available because the DEBUT study is still ongoing. The dataset may be available upon request after 2022 once the main DEBUT study is completed as described by the National Institute of Health.

## Abbreviations

ASCRS, American Society of Colorectal Surgeons; AUC, area under the curve; CT, computed tomography; DEBUT, Diverticulitis Evaluation of Patient Burden, Utilization, and Trajectory; DV-QOL, diverticulitis quality of life instrument; ED, emergency department; GI, gastrointestinal; GIQLI, Gastrointestinal Quality of Life Index; HRQoL, health-related quality of life; IQR, interquartile range; MCID, minimum clinically important difference; PASS, patient acceptable symptom state; PRO, patient-reported outcome; PROMIS, Patient-Reported Outcomes Measurement Information System; Q_max_, quarter with the highest DV-QOL score; ROC, the receiver operating characteristic; SD, standard deviation

## Ethics approval and consent to participate

This study was approved by the University of Washington Institutional Review Board (IRB) (STUDY00003708), the for the following study sites: University of Washington Medical Center, Harborview Medical Center, Northwest Hospital, Legacy Health, and Valley Medical Center. Approval was also attained for the Ronald Reagan UCLA Medical Center and Skagit Regional Health from the UCLA IRB (#16-000599) and Skagit Regional Health IRB (DEBUT study), respectively. Patient consent was given in writing, online, or in person with a research coordinator.

## References

1. Peery AF, Crockett SD, Murphy CC, et al (2019) Burden and cost of gastrointestinal, liver, and pancreatic diseases in the United States: update 2018. Gastroenterology 156(1):254-272. e11.

2. Eglinton T, Nguyen T, Raniga S, et al (2010) Patterns of recurrence in patients with acute diverticulitis. Br J Surg 97(6):952–957.

3. Rafferty J, Shellito P, Hyman NH, et al (2006) Practice parameters for sigmoid diverticulitis. Dis Colon Rectum 49(7):939–944.

4. Feingold D, Steele SR, Lee S, et al (2014) Practice parameters for the treatment of sigmoid diverticulitis. Dis Colon Rectum 57(3):284–294.

5. Strate LL, Modi R, Cohen E, et al (2012) Diverticular disease as a chronic illness: evolving epidemiologic and clinical insights. Am J Gastroenterol 107(10):1486–1493.

6. Spiegel BM, Reid MW, Bolus R, et al (2015) Development and validation of a disease-targeted quality of life instrument for chronic diverticular disease: the DV-QOL. Qual Life Res 24(1):163–179.

7. Eypasche E, Williams JI, Wooddauphinee S, et al (1995) Gastrointestinal quality-of-life index - development, validation and application of a new instrument. Br J Surg 82(2):216–222.

8. Patrick DL and Deyo RA (1989) Generic and disease-specific measures in assessing health-status and quality of life. Med Care 27(3):S21–S232.

9. Stratford PW, Binkley JM, Riddle DL, Guyatt GH (1998) Sensitivity to change of the roland-morris back pain questionnaire: Part 1. Phys Ther 78(11):1186–1196.

10. Tubach F, Ravaud P, Baron G, et al (2005) Evaluation of clinically relevant states in patient reported outcomes in knee and hip osteoarthritis: The patient acceptable symptom state. Ann Rheum Dis 64(1):34–37.

11. Comparative Effective Research Translation Network. Diverticulitis evaluation of patient burden, utilization and trajectory (DEBUT) study. https://www.becertain.org/debut-diverticulitis-study. Accessed 14 February 2020.

12. Copay AG, Subach BR, Glassman SD, et al (2007) Understanding the minimum clinically important difference: a review of concepts and methods. Spine J 7(5):541–546.

13. Norman GR, Sloan JA, Wyrwich KW (2004) The truly remarkable universality of half a standard deviation: Confirmation through another look. Expert Rev Pharmacoecon Outcomes Res 4(5):581–585.

14. Norman GR, Sloan JA, Wyrwich KW (2003) Interpretation of changes in health-related quality of life - the remarkable universality of half a standard deviation. Med Care 41(5):582–592.

15. Cella D, Riley W, Stone A, et al (2010) The patient-reported outcomes measurement information system (PROMIS) developed and tested its first wave of adult self-reported health outcome item banks: 2005-2008. J Clin Epidemiol 63(11):1179–1194.

16. Hays RD, Bjorner JB, Revicki DA, et al (2009) Development of physical and mental health summary scores from the patient-reported outcomes measurement information system (PROMIS) global items. Qual Life Res 18(7):873–880.

17. Deyo RA (1984) Pitfalls in measuring the health-status of Mexican-Americans: comparative validity of the English and Spanish Sickness Impact Profile. Am J Public Health 74(6):569–573.

18. Reilly MC, Zbrozek AS, Dukes EM (1993) The validity and reproducibility of a work productivity and activity impairment instrument. Pharmacoeconomics 4(5):353–365.

19. Spiegel BMR, Hays RD, Bolus R, et al (2014) Development of the NIH patient-reported outcomes measurement information system (PROMIS) gastrointestinal symptom scales. Am J Gastroenterol 109(11):1804–1814.

20. Mackenzie CR, Charlson ME, Digioia D, et al (1986) Can the Sickness Impact Profile measure change? An example of scale assessment. J Chronic Dis 39(6):429–438.

